# TractLearn: a geodesic learning framework for quantitative analysis of brain bundles

**DOI:** 10.1101/2020.05.27.20113027

**Authors:** Arnaud Attyé, Félix Renard, Monica Baciu, Elise Roger, Laurent Lamalle, Patrick Dehail, Hélène Cassoudesalle, Fernando Calamante

**Author notes:** Corresponding author: Arnaud Attyé, Permanent address: CNRS LPNC UMR 5105, University of Grenoble Alpes, Grenoble, France. Declarations of interest: none for the present work.

## Abstract

Deep learning-based convolutional neural networks have recently proved their efficiency in providing fast segmentation of major brain fascicles structures, based on diffusion-weighted imaging. The quantitative analysis of brain fascicles then relies on metrics either coming from the tractography process itself or from each voxel along the bundle.

Statistical detection of abnormal voxels in the context of disease usually relies on univariate and multivariate statistics models, such as the General Linear Model (GLM). Yet in the case of high-dimensional low sample size data, the GLM often implies high standard deviation range in controls due to anatomical variability, despite the commonly used smoothing process. This can lead to difficulties to detect subtle quantitative alterations from a brain bundle at the voxel scale.

Here we introduce *TractLearn*, a unified framework for brain fascicles quantitative analyses by using geodesic learning as a data-driven learning task. *TractLearn* allows a mapping between the image high-dimensional domain and the reduced latent space of brain fascicles using a Riemannian approach.

We illustrate the robustness of this method on a healthy population with test-retest acquisition of multi-shell diffusion MRI data, demonstrating that it is possible to separately study the global effect due to different MRI sessions from the effect of local bundle alterations. We have then tested the efficiency of our algorithm on a sample of 5 age-matched subjects referred with mild traumatic brain injury.

Our contributions are to propose an algorithm based on:

1/ A manifold approach to capture controls variability as standard reference instead of an atlas approach based on a Euclidean mean

2/ The ability to detect global variation of voxels quantitative values, which means that all the voxels interaction in a structure are considered rather than analyzing each voxel independently.

With this regard, *TractLearn* is a ready-to-use algorithm for precision medicine.

**KEY POINT:**

- We provide a statistical test taking into account the interaction between voxels
- We propose to use a Riemaniann manifold as reference instead of a Euclidean mean
- We demonstrate the usefulness and reliability of the track-weighted contrast

## I. INTRODUCTION

A streamline data-set generated from diffusion MRI provides a wealth of information regarding structural connectivity between brain regions in medicine and neurosciences. An elegant approach to segment brain bundles has recently been proposed, TractSeg, based on a combination of three convolutional neural networks (Wasserthal et al., 2019). The first network proposes to segment major bundles using a voxel-wise binary classification to discern tract and non-tract voxels. The second network allows to learn the start and end regions of each brain fascicle, while the last one computes tract orientations mapping to obtain a single 3D peak vector per voxel. This architecture is then employed to start a probabilistic tracking algorithm for 72 brain bundles, tested in various acquisition conditions and on a few disease models. We could consider this segmentation process as a first step to reduce the dimensionality of data coming from a whole-brain diffusion MRI acquisition.

Usually, group study statistical analyses rely on the General Linear Model (GLM), a standard tool based on Gaussian distribution assumptions that returns for each voxel the mean of the control group. The subsequent step is either t-tests in case of group vs group studies or Z-score in case of the comparison between one individual versus a group; this is usually done analyzing each voxel independently. In addition, precision medicine requires to have algorithms suitable to manage high dimensional low sample size data (HDLSS). A potential limitation of HDLSS is the difficulties to capture all the physiological variation of MRI contrasts using a Euclidean mean, leading to high standard-deviation values (see supplementary figures 1 and 2, for a simulated illustrative example). It can theoretically limit the possibility to distinguish bundle subtle quantitative alterations.

One way to address these two limitations is by manifold learning. Manifold learning is a class of machine learning methods that is gaining success and attracting interest, allowing reconstruction of a manifold sub-space to represent, understand and visualize degrees of freedom of complex quantitative data (Tenenbaum et al., 2000). It is classically used before deep learning algorithms to identify outliers, for example to increase the robustness of images reconstructions (Zhu et al., 2018).

While classical deep learning algorithms allow to segment, identify, and classify imaging features based on big data, a manifold framework usually requires multiple quantitative biomarkers per region of interest to estimate distances between subjects, while still remaining applicable to studies with a low number of imaging data. *UMAP* (Uniform Manifold Approximation and Projection) is a recent nonlinear manifold learning technique for dimensionality reduction, which is constructed from a theoretical framework based on Riemannian geometry and algebraic topology (McInnes et al., 2018). The result is a practical scalable algorithm that applies to real world data.

The information contained in the streamline tractograms can be exploited to generate images with different contrasts using the track-weighted imaging (TWI) framework (Calamante, 2017; Calamante et al., 2012). The TWI contrast has proved to be useful for clinical care in providing fast identification of major alterations in brain fascicles (e.g. tumor proliferation (Barajas et al., 2013) or demyelination (Lyksborg et al., 2014). Yet, it might be also relevant to highlight subtler quantitative alterations at the global level.

As the quantitative and reliability aspects of the TWI contrast has only been investigated in the context of whole brain tractography (Calamante et al., 2015; Willats et al., 2014), new test-retest data manifold learning analysis would be interesting to compare diseased individuals to a healthy cohort within a Riemannian framework. In other words, we raise the hypothesis that manifold learning can be used to detect local abnormalities in white matter bundles, and that the detected abnormalities will be robust to the healthy control data set used (i.e. test/retest data) as reference.

Here, we propose a 3-step method to obtain a fast-quantitative analysis of brain bundles, through various quantitative metrics. Firstly, we will reduce the dimensionality of all voxels contained in each bundle, to have one point per bundle and subject in a manifold subspace. Secondly, we will learn the manifold from the test and the retest session of our healthy controls independently (Tilquin et al., 2019) before projection of the five trauma patients onto the learned manifolds. Finally, we propose to apply Z-scores to the projection of a new subject onto the learned manifolds (from the test and retest sessions) before calculating inter-session agreement to detect local abnormalities.

## II. METHODS

### 1. Data and code availability statements

The code was based on the *UMAP* algorithm (McInnes et al., 2018), freely available on https://github.com/lmcinnes/umap and on the TractSeg pre-trained DWI algorithm, as openly available at https://github.com/MIC-DKFZ/TractSeg (Wasserthal et al., 2018a). The code for *TractLearn* will be uploaded on Github on acceptance of a final version of the manuscript.

### 2. Data acquisition

20 healthy males (mean age 20.9 [SD 3.3] years) were recruited for the study. Informed written consent was obtained from all subjects in accordance with ethical approval from the local human research ethics committee of Bordeaux university (France, IRB number 2016-A00765-46). MRI data were acquired on a 3 T Siemens Prisma scanner using a 48-channel head coil. High angular resolution diffusion imaging (HARDI) datasets were acquired for each subject over two sessions with a delay of 12 months. Five age-matched subjects referred with mild traumatic brain injury (mTBI) were also included with the same MR protocol and ethical statements, for one unique MR session acquired between the time of the test and retest session for the controls.

Diffusion-weighted imaging (DWI): single-shot spin-echo sequence; TE/TR: 80/4700 ms; voxel size: 1.5 mm isotropic; 99 slices; multiband factor: 3; scan time: 7.5 min. In addition to 10 non-DWI volumes (which were averaged), 3 DWI shells were acquired, each with a different diffusion weighting and a unique set of diffusion-weighted directions. The set of directions was independently generated for each shell by electrostatic repulsion, as follow: directions/b-value (in s/mm^2^) = 60/2000, 15/800, and 10/300.

In order to allow pre-processing minimization of distortion artefacts, two additional b = 0 s/mm^2^ images were acquired before each HARDI acquisition, with identical imaging parameters as above, but one had its phase encoding reversed to allow for susceptibility distortion correction (Holland et al., 2010).

T1-weighted anatomical images were acquired using the three-dimensional magnetization-prepared rapid gradient echo (3D MPRAGE) sequence (Mugler and Brookeman, 1990) with the following parameters: 256 × 256 × 192 matrix; 0.9 mm isotropic resolution; TE 2.6 ms; inversion time TI 900 ms; TR 1900 ms; flip angle 9°. Susceptibility-weighted imaging and 3D-FLAIR sequences were also acquired.

### 3. Data pre-processing

DWI preprocessing included denoising of data (Veraart et al., 2016), eddy current correction and motion correction (Andersson et al., 2003), bias field and Gibbs artefacts corrections (Tustison et al., 2010), and up-sampling DWI spatial resolution in all three dimensions using cubic b-spline interpolation, to a voxel size of 1 mm isotropic (Raffelt et al., 2012a). We have estimated fiber orientation distributions (FODs) using the Constrained Spherical Deconvolution (CSD) model (Tournier et al., 2007) using group response function (RF); in particular, we used the multi-shell 3-tissue CSD variant (Jeurissen et al., 2014). We derived the SH peaks from the FOD maps. Spatial correspondence was achieved by first generating a group-specific population template with an iterative registration and averaging approach (Raffelt et al., 2011) using FOD images from 45 MR scans (5 mTBI, 20 healthy control subjects acquired in a test session, and 20 healthy control subjects acquired in a retest session). Each subject’s FOD image was then registered to the template via a FOD-guided non-linear registration (Raffelt et al., 2011, 2012a).

All preprocessing steps were conducted using commands either implemented within MRtrix3 (Tournier et al., 2019) (www.mrtrix.org), or using MRtrix3 scripts that interfaced with external software packages.

### 4. TractSeg Deep learning bundle specific tractography

We have used the TractSeg pre-trained algorithm to automatically identify 72 white matter bundles in each subject. Briefly, authors have proposed a custom probabilistic tracking algorithm that samples from a Gaussian distribution with fixed standard deviation centered on each spherical harmonic peak. They have used three convolutional neural networks (tract segmentation, start/end region segmentation and tract orientation mapping), all based on U-Net (Ronneberger et al., 2015) that receives as input the fiber orientation distribution function (FOD) peaks.

Importantly, for our application, we have warped all the resulting TractSeg tracks from each individual space into the common FOD template space, before the subsequent steps of our analysis.

### 5. Estimating track-weighted imaging

Track-weighted imaging (TWI) represents information related to the streamlines in a 3D image format (which can have super-resolution properties); for example, TWI maps can be generated from various properties of the streamlines themselves, such as number of streamlines in each voxel or their average length (Calamante, 2016; Pannek et al., 2011). Alternatively, TWI maps can also be computed from track-weighted version (e.g. an average along the track) of the contrast of an associated parameter (e.g. a fractional anisotropy map).

As the manifold approach requires an accurate voxel to voxel matching between subjects, we first converted each track file into TWI maps based on the number of streamlines (also known as track-density imaging (TDI) maps (Calamante et al., 2010)). The objective was to crop the bundle masks, only keeping the voxels with top 80% TDI values (a value that was empirically chosen). This allows to decrease potential misregistration of brain bundles between subjects by removing the smallest cortical terminations, which can be highly variable at the group level.

Secondly, we computed TWI maps using a Gaussian-smoothed kernel for computing the parameter along each streamline; a full-width half-maximum kernel was set to 8 mm, as an empirical compromise between spatial smoothing (for increase signal to noise) and along-tract spatial blurring (to be able to detect localized effects) (Calamante, 2017; Willats et al., 2014). In particular, for the results presented here, we considered two types of TWI maps: TWI based on the Fractional anisotropy (TW-FA), and TWI based on the amplitude of the FOD (TW-FOD) along the direction of the track (Willats et al., 2014). Finally, for comparison purpose of not using the TWI approach, we have extracted from each bundle the voxel values from the FA and an FOD related map. To construct the FOD-related map (Raffelt et al., 2012b), we used the *afdconnectivity* command in MRtrix, which provides an estimate of the fibre volume of the pathway of interest at each voxel.^1^ Figure 1 shows an overview of the workflow of the analysis.

**Figure 1:**
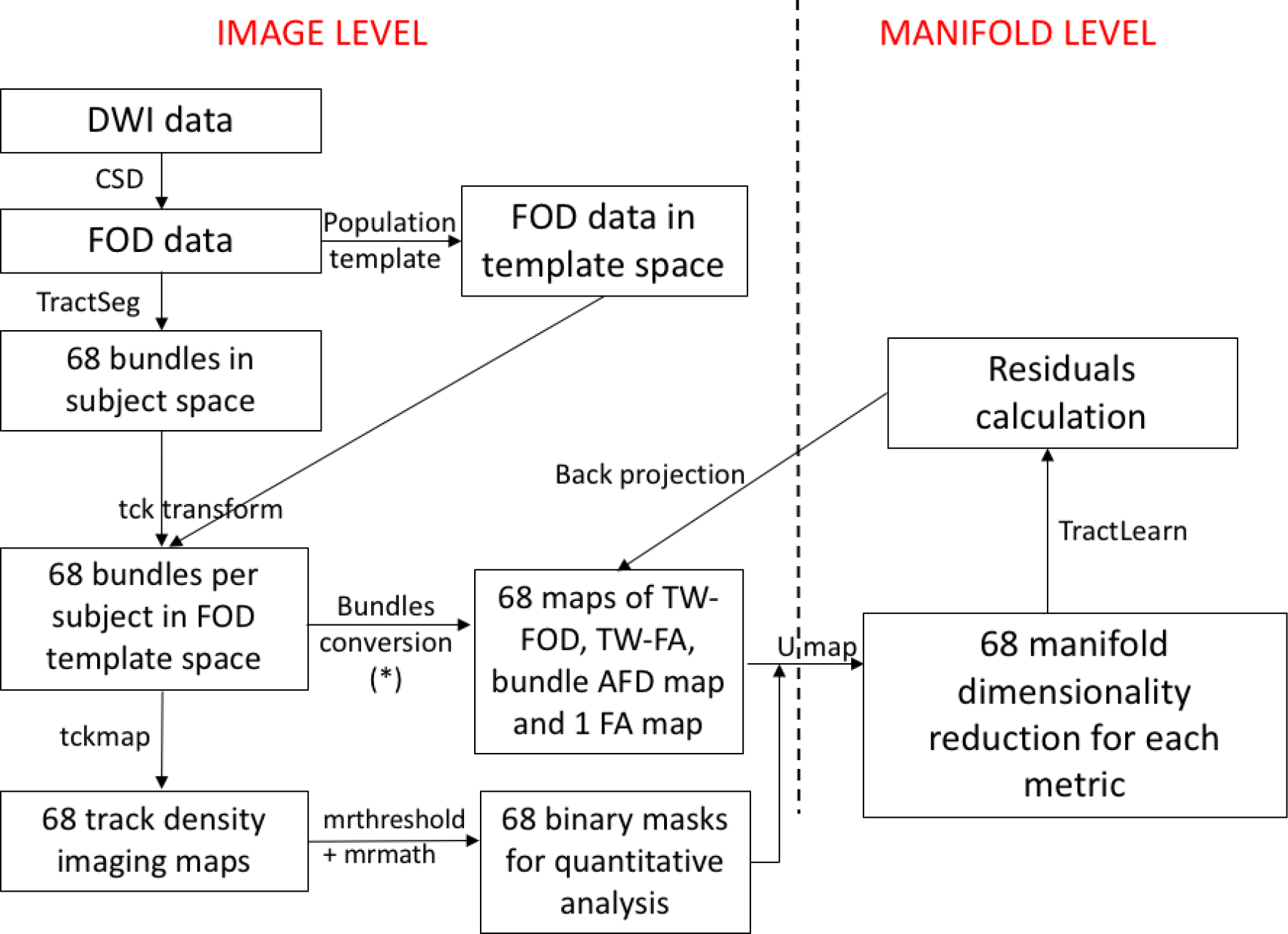
Overview of the workflow put forward in this work. The bundle-to-image conversion (*) was obtained in MRtrix either using *tckmap* (for TW-FOD and TWI-FA contrasts) or *afdconnectivity* (for the AFD-related contrast). The FA contrast was obtained directly from the FA map. The number of bundles is explained at the beginning of the results part.

**Figure 2:**
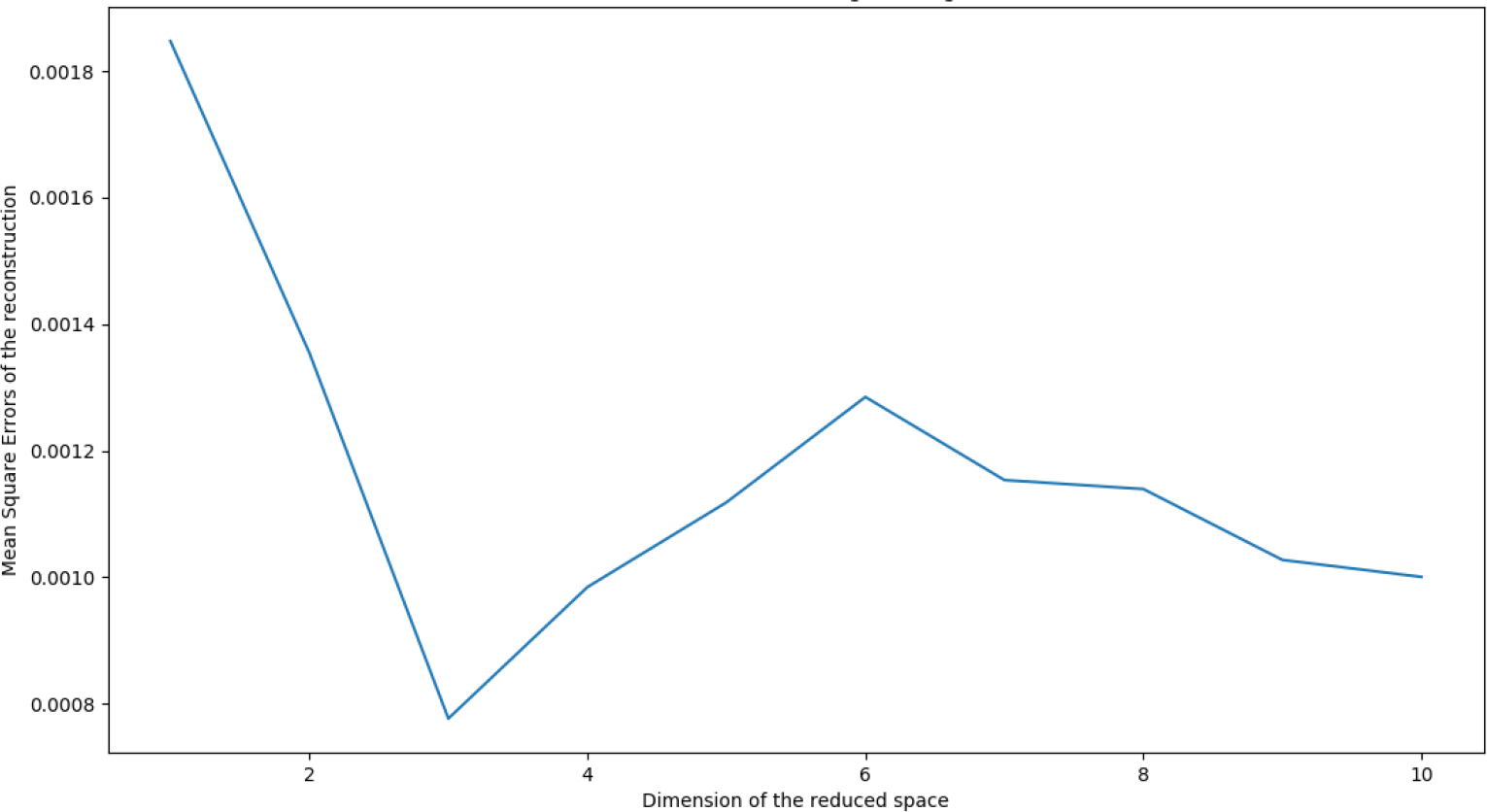
Example of the right cingulum, showing a minimum mean square error of the reconstructions for a dimension equal to 3. Each point of the curve represents a single execution of the *TractLearn* algorithm with a particular regularization: the x- and y-axes positions are determined by the dimension of the reduced space and mean square errors of the reconstruction, respectively.

### 6. Modeling brain fascicles quantitative values with manifold learning

We used a strategy similar to that developed in (Tilquin et al., 2019) to localize an abnormality among *TractSeg* bundles. Note that while *TractLearn* uses a Riemannian framework to estimate the distance between all metrics contained in brain bundles, the statistics in the low-dimensional reduced manifold space (for example the distance between a point corresponding to a patient and a point corresponding to a healthy control in the manifold subspace) are then performed following classical Euclidean metrics. Here we assume that the residuals in the reduced subspaces follow a classic Gaussian law (Titsias and Lawrence, 2004).

*TractLeam* can be summarized as follow:

i. Low-dimensional reduction to convert collection of voxels quantitative values from each bundle into a unique point in a manifold subspace. To test the effect of dimension reduction on the MRI data, we performed optimization-based methods. We have chosen the number of dimension by repeating the reduction process using an increasing number of dimensions and evaluating whether incorporating more components achieves a significantly lower value of the loss function that the method minimizes (Nguyen and Holmes, 2019).
ii. Here we have used *UMAP* (McInnes et al., 2018). *UMAP* is a manifold learning approach, where the degrees of freedom of the data are captured by the latent variables. Importantly, the structure of points in the latent space (the reduced space) mimics the structure of data in the original space. Interpoint distances in the reduced space reproduce as much as possible interpoint distances in the original space; Euclidean and geodesic distances are respectively used. We investigate two issues: (a) interpreting the latent variables, and (b) determining the effect a change in the latent variables incurs in the MRI space, i.e. the corresponding changes in identified brain bundles voxels.
iii. We learn the manifold based on the data from the healthy controls: *Y=f(x)*+*ε*. *Y* being healthy control data in real space (i.e. quantitative values extracted from each *TractSeg* bundle), *x* the corresponding point in the reduced space, and *ε* the residuals; *f* will be the regression equation between the reduced space and the real space. The projection *f* of a new subject will correspond to the image closest to that of the tested subject, while belonging to the manifold of the healthy controls. It is worthwhile to note that, in the Riemaniann framework, *f(x)* represents the local mean value (ie. the mean value of the closest healthy subject) of the tested metric in the reduced space (Titsias and Lawrence, 2004). For a comparison purpose, the GLM was also used: Y=Euclidean Mean + *ε_GLM._* In other words, as the Riemaniann atlas has been built to capture the maximum variability, a new subject projected onto the learned manifold (for example the projection of one mTBI patient) will be assigned using a local average to the closest individuals of the 20 healthy controls using the Nadaraya-Watson kernel estimator for high dimensional non parametric regression (Conn and Li, 2017). The selected healthy controls will be those who have closest quantitative values from the tested subject. Consequently, it is expected that f(x) will be generally less than the Euclidean mean, allowing *ε* to be more representative of the pathological changes in the Riemaniann framework. Indeed, a classical Euclidean *ε_GLM_* does represent both potential pathological changes and anatomical variability (i.e. the standard deviation of the mean) (Tilquin et al., 2019; Vik et al., 2007). The choice to work on a manifold (either Riemaniann in our case, or using a more classical linear principal component analysis) is to shorten the standard deviation value so that *ε* mainly represents the potential disease effect. A visual comparison of standard deviation between the GLM and *TractLearn* is provided in supplementary figure 1.
iv. To ensure robustness in our dataset, *ε* was estimated using a leave-one-out (LOO) strategy, by randomly selecting inter and intra-individual distances in the manifold from the test session. We consider that the residual *ε* is representative of the abnormalities present in a new subject when it is greater than the model variability learned during the LOO on the healthy control group. A total of 1360 permutation tests were computed in the control group using the LOO. At this step, we have obtained *ε* estimation in all controls allowing a comparison with the *ε* estimated from the 5 mTBI patients. We make the assumption that £ follows a multivariate Gaussian distribution with a standard deviation that varies across the voxels (Tilquin et al., 2019; Vik et al., 2007).
v. We finally identified altered voxels in each fascicle in real space by applying a Z-score. A Bonferroni correction was performed for the testing of a mean of n voxels tests (dividing the p-value by a factor of n). The number of n voxels corresponds to all the voxels which are contained in each TractSeg generated bundles (for example, a bundle of 20,000 voxels was analyzed by dividing the p-value by a factor of 20,000). A Bonferroni-corrected significance threshold of 0.05 was used. Furthermore, we have also generated a synthetic lesion on corpus callosum 3 to illustrate the difference between Z scores in our Riemaniann frameworks and using the GLM (supplementary figure 2).

### 7. Test-Retest statistics

We have used three strategies to estimate the test-retest reliability of each voxel metric in the TractSeg bundles.

Firstly, we have calculated intra-class correlation coefficients (ICC) for the different metrics by comparing the mean values of all voxels contained in one bundle between the test and the retest session. It is a classic approach in which a single-score ICC is based on a two-way model. A paired t-test was used to compare TW-FA and FA from one side, TW-FOD and AFD from the other side.

Secondly, we have compared the distance between the test and the retest session in the reduced space for each bundle with the distance between the controls and each patient, as illustrated figure 3.

**Figure 3:**
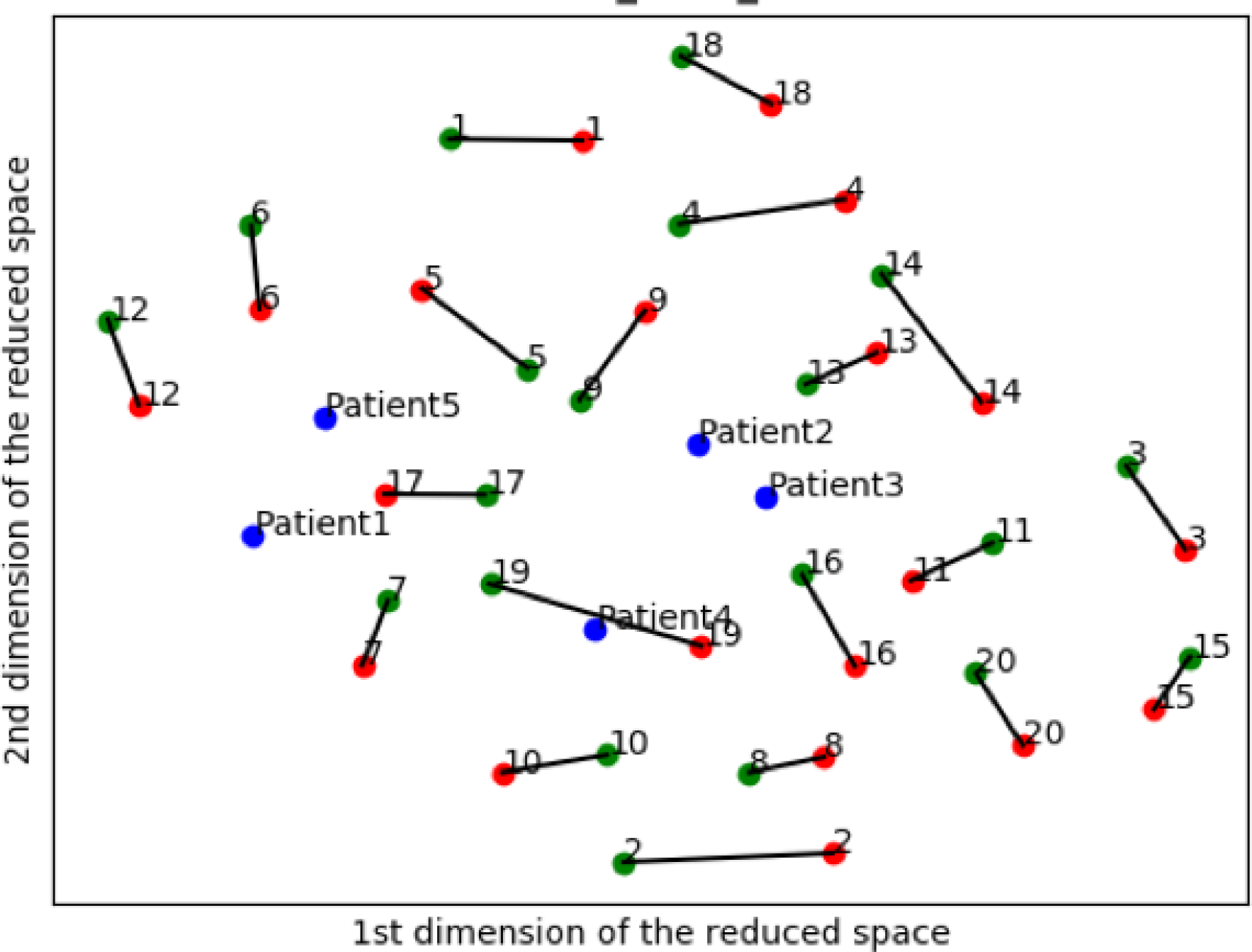
2D reduced space of left thalamo-prefrontal bundles. The units of each axes are usually arbitrary in dimensionality reduction. Here the TW-FOD metrics are provided as input for *TractLearn*. In this framework, a point represents a collection of TW-FOD values (one per voxel in a given bundle) in the reduced subspace. The farther two points are from each other, the more different the quantitative metric values are. Healthy controls are numerated from 1 to 20, with green dots representing the test session, and red dots representing the retest session 12 months later; the mTBI patients are represented by the five blue dots. For most subjects, red and green dots are in close vicinity in the manifold, with the black lines representing the “MRI inter-session effect”. The patients bundle collection of metrics here are located among those of the controls, suggesting that there is no or low global difference between controls and patients in this randomly selected bundle.

Finally, we have evaluated the number of injured bundles in the 5 mTBI patients depending on the choice of session for training the manifold space, i.e. when the test (or retest) session was used to compute the learned manifold of the healthy group. To allow loss functions comparisons (i.e. to statistically compare the inaccuracy of predictions in detecting the number of lesions between the test and retest session), we have considered lesions with at least 5 altered voxels per bundle.

## III. RESULTS

### 1. TractSeg and metrics reliability

All but 2 (fornix and anterior commissure) of the 72 brain bundles were successfully reconstructed in all subjects using *TractSeg*. As the anterior commissure and the fornix were not robustly reconstructed in all subjects, they have been excluded from further quantitative analyses. We have also chosen to study the corpus callosum through the seven subparts identified by *TractSeg* (rather than as a single anatomical structure), to help with the localization of potential lesions. Our analysis is therefore restricted to the remaining 68 bundles (all *TractSeg* bundles but left fornix, right fornix, anterior commissure, and non-segmented corpus callosum).

The names and abbreviations of each brain bundle are as follow (Wasserthal et al., 2018b): Arcuate fascicle (AF), Anterior thalamic radiation (ATR), Corpus callosum (Rostrum (CC 1), Genu (CC 2), Rostral body (CC 3), Anterior midbody (CC 4), Posterior midbody (CC 5), Isthmus (CC 6), Splenium (CC 7)), Cingulum (CG), Corticospinal tract (CST), Middle longitudinal fascicle (MLF), Fronto-pontine tract (FPT), Inferior cerebellar peduncle (ICP), Inferior occipito-frontal fascicle (IFO), Inferior longitudinal fascicle (ILF), Middle cerebellar peduncle (MCP), Optic radiation (OR), Parieto-occipital pontine (POPT), Superior cerebellar peduncle (SCP), Superior longitudinal fascicle I (SLF I), Superior longitudinal fascicle II (SLF II), Superior longitudinal fascicle III (SLF III), Superior thalamic radiation (STR), Uncinate fascicle (UF), Thalamo-prefrontal (T PREF), Thalamo-premotor (T PREM), Thalamo-precentral (T PREC), Thalamo-postcentral (T POSTC), Thalamo-parietal (T PAR), Thalamo-occipital (T OCC), Striato-fronto-orbital (ST FO), Striato-prefrontal (ST PREF), Striato-premotor (ST PREM), Striato-precentral (ST PREC), Striato-postcentral (ST POSTC), Striato-parietal (ST PAR), Striato-occipital (ST OCC). Note: besides MCP and the 7 CC subdivisions, all other bundles were present in each hemisphere.

The MRI inter-session effect of the metrics measured in the *TractSeg* bundles was firstly studied by the ICC coefficients. ICC was on average 0.89, 0.88, 0.85 and 0.84 for TW-FA, FA, TW-FOD and bundle AFD, respectively; individual bundle ICC values are shown in Table 1.

**Table 1:** ICC coefficients for all 4 metrics mean value in each TractSeg bundle.

|  | TW-FA | FA | TW-FOD | Bundle<br>AFD |  | TW-FA | FA | TW-FOD | Bundle<br>AFD |
| --- | --- | --- | --- | --- | --- | --- | --- | --- | --- |
| AF_left | 0,93 | 0,92 | 0,97 | 0,94 | SLF_II_left | 0,81 | 0,78 | 0,84 | 0,81 |
| AF_right | 0,95 | 0,94 | 0,96 | 0,94 | SLF_II_right | 0,9 | 0,87 | 0,92 | 0,89 |
| ATR_left | 0,9 | 0,9 | 0,82 | 0,84 | SLF_I_left | 0,8 | 0,78 | 0,7 | 0,77 |
| ATR_right | 0,89 | 0,9 | 0,78 | 0,78 | SLF_I_right | 0,87 | 0,85 | 0,8 | 0,57 |
| CC_1 | 0,82 | 0,81 | 0,81 | 0,78 | STR_left | 0,91 | 0,91 | 0,73 | 0,88 |
| CC_2 | 0,91 | 0,89 | 0,87 | 0,87 | STR_right | 0,91 | 0,9 | 0,82 | 0,87 |
| CC_3 | 0,81 | 0,77 | 0,76 | 0,65 | ST_FO_left | 0,93 | 0,93 | 0,84 | 0,87 |
| CC_4 | 0,82 | 0,81 | 0,83 | 0,79 | ST_FO_right | 0,8 | 0,78 | 0,71 | 0,52 |
| CC_5 | 0,92 | 0,93 | 0,91 | 0,97 | ST_OCC_left | 0,95 | 0,94 | 0,96 | 0,92 |
| CC_6 | 0,9 | 0,91 | 0,9 | 0,89 | ST_OCC_right | 0,91 | 0,91 | 0,97 | 0,94 |
| CC_7 | 0,9 | 0,91 | 0,81 | 0,7 | ST_PAR_left | 0,91 | 0,9 | 0,92 | 0,92 |
| CG_left | 0,88 | 0,86 | 0,92 | 0,85 | ST_PAR_right | 0,89 | 0,88 | 0,95 | 0,94 |
| CG_right | 0,86 | 0,85 | 0,93 | 0,85 | ST_POSTC_left | 0,86 | 0,83 | 0,73 | 0,75 |
| CST_left | 0,96 | 0,96 | 0,92 | 0,92 | ST_POSTC_right | 0,9 | 0,89 | 0,85 | 0,81 |
| CST_right | 0,85 | 0,85 | 0,87 | 0,79 | ST_PREC_left | 0,94 | 0,94 | 0,89 | 0,93 |
| FPT_left | 0,93 | 0,92 | 0,9 | 0,88 | ST_PREC_right | 0,94 | 0,94 | 0,91 | 0,92 |
| FPT_right | 0,95 | 0,95 | 0,94 | 0,9 | ST_PREF_left | 0,79 | 0,74 | 0,66 | 0,55 |
| ICP_left | 0,79 | 0,79 | 0,67 | 0,66 | ST_PREF_right | 0,91 | 0,9 | 0,89 | 0,86 |
| ICP_right | 0,83 | 0,83 | 0,82 | 0,86 | ST_PREM_left | 0,84 | 0,71 | 0,77 | 0,64 |
| IFO_left | 0,94 | 0,94 | 0,94 | 0,94 | ST_PREM_right | 0,93 | 0,94 | 0,78 | 0,88 |
| IFO_right | 0,87 | 0,87 | 0,88 | 0,84 | T_OCC_left | 0,96 | 0,96 | 0,96 | 0,93 |
| ILF_left | 0,92 | 0,92 | 0,91 | 0,9 | T_OCC_right | 0,84 | 0,86 | 0,88 | 0,87 |
| ILF_right | 0,75 | 0,75 | 0,62 | 0,8 | T_PAR_left | 0,89 | 0,88 | 0,78 | 0,89 |
| MCP | 0,9 | 0,9 | 0,87 | 0,87 | T_PAR_right | 0,85 | 0,85 | 0,81 | 0,85 |
| MLF_left | 0,91 | 0,88 | 0,9 | 0,85 | T_POSTC_left | 0,91 | 0,89 | 0,75 | 0,88 |
| MLF_right | 0,95 | 0,95 | 0,86 | 0,93 | T_POSTC_right | 0,96 | 0,96 | 0,73 | 0,86 |
| OR_left | 0,97 | 0,97 | 0,96 | 0,93 | T_PREC_left | 0,91 | 0,91 | 0,67 | 0,85 |
| OR_right | 0,8 | 0,79 | 0,87 | 0,89 | T_PREC_right | 0,93 | 0,92 | 0,87 | 0,9 |
| POPT_left | 0,92 | 0,93 | 0,91 | 0,92 | T_PREF_left | 0,9 | 0,9 | 0,85 | 0,87 |
| POPT_right | 0,95 | 0,94 | 0,9 | 0,9 | T_PREF_right | 0,95 | 0,95 | 0,9 | 0,88 |
| SCP_left | 0,84 | 0,84 | 0,84 | 0,89 | T_PREM_left | 0,88 | 0,89 | 0,93 | 0,93 |
| SCP_right | 0,84 | 0,82 | 0,82 | 0,84 | T_PREM_right | 0,94 | 0,94 | 0,77 | 0,87 |
| SLF_III_left | 0,91 | 0,89 | 0,86 | 0,93 | UF_left | 0,84 | 0,8 | 0,9 | 0,88 |
| SLF_III_right | 0,89 | 0,86 | 0,89 | 0,89 | UF_right | 0,84 | 0,81 | 0,88 | 0,74 |

The t-tests, as paired per bundle, showed significantly higher ICC values for TW-FA in comparison to FA (p=0.00014) but not for TW-FOD as compared with Bundle AFD (p=0.86).

### 2. TractLearn

We have firstly determined that the dimension with the minimum mean square error was equal to 3 for all 68 bundles. For visualization purpose, we have chosen to graphically present the reduced space as 2-dimensional plots, though all the residual estimation was based on a 3-dimensional reduction of each brain bundle.

The inter-session effect was then directly studied in the manifold. Using *TractLearn*, the distance in the geodesic space for each bundle was significantly shorter between the test and the retest procedure (intra-subject) than between two different subjects (p<0.001) (e.g. see example in Figure 3).

In all patients, the bundle AFD metric found the highest number of bundles with altered voxels. When taking into account the results from the manifold based on the test and the manifold based on the retest lesion, patient number 5 presented with the higher number of injured bundles (including corpus callosum parts 4 and 7, right thalamo-occipital bundle, right striato-occipital bundle, right optic radiation and middle cerebellar peduncle), with some of them including at least 20 altered voxels. Patient 2 showed no bundle with more than 10 altered voxels for any metrics (Figure 5) – see Supplementary Material for the corresponding results of the other three patients.

**Figure 4:**
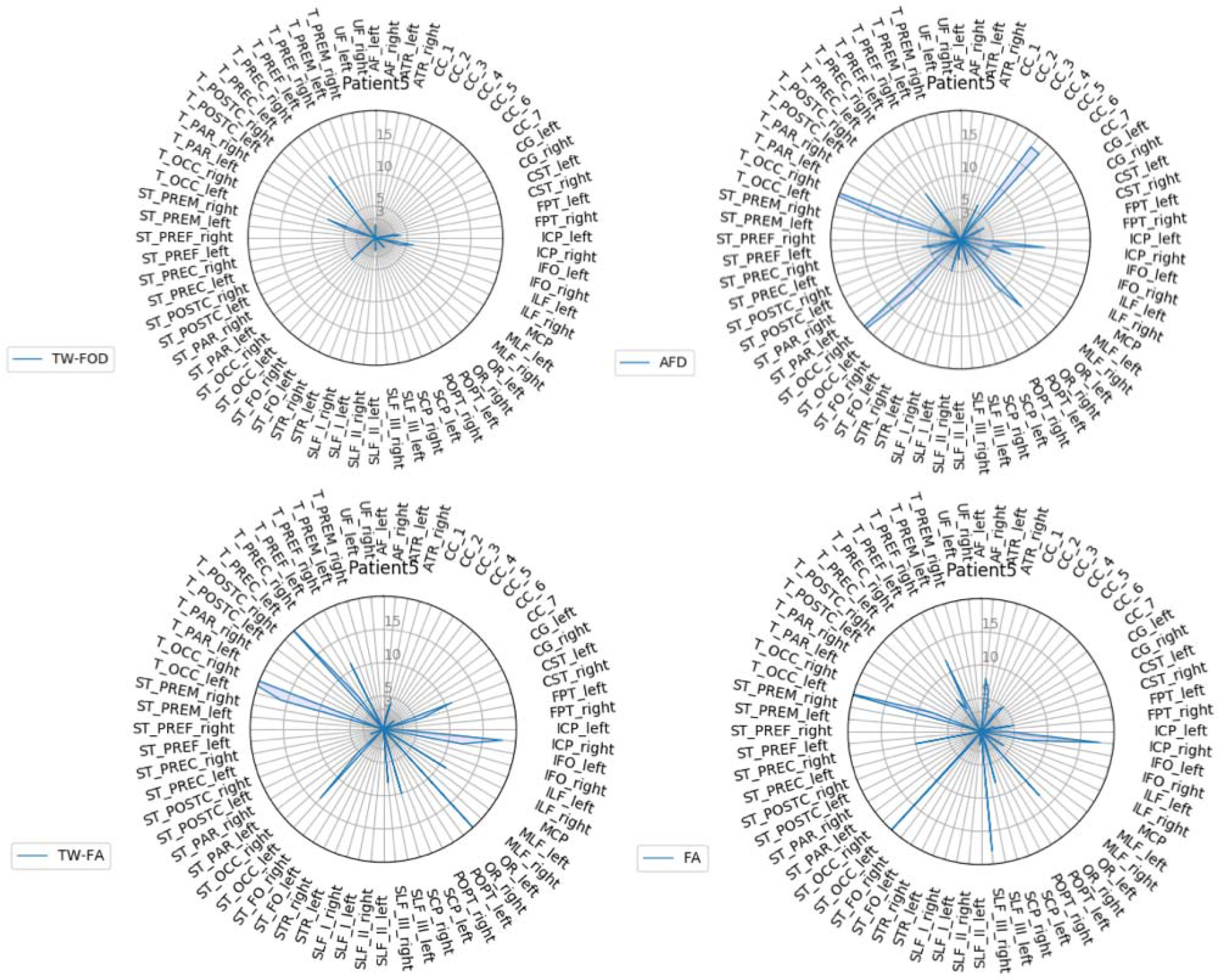
Radar plots of *TractLearn* results for patient 5 showing the number of voxels presenting with significant altered Z scores in each bundle, as identified using *TractLearn* based on session 1.

**Figure 5:**
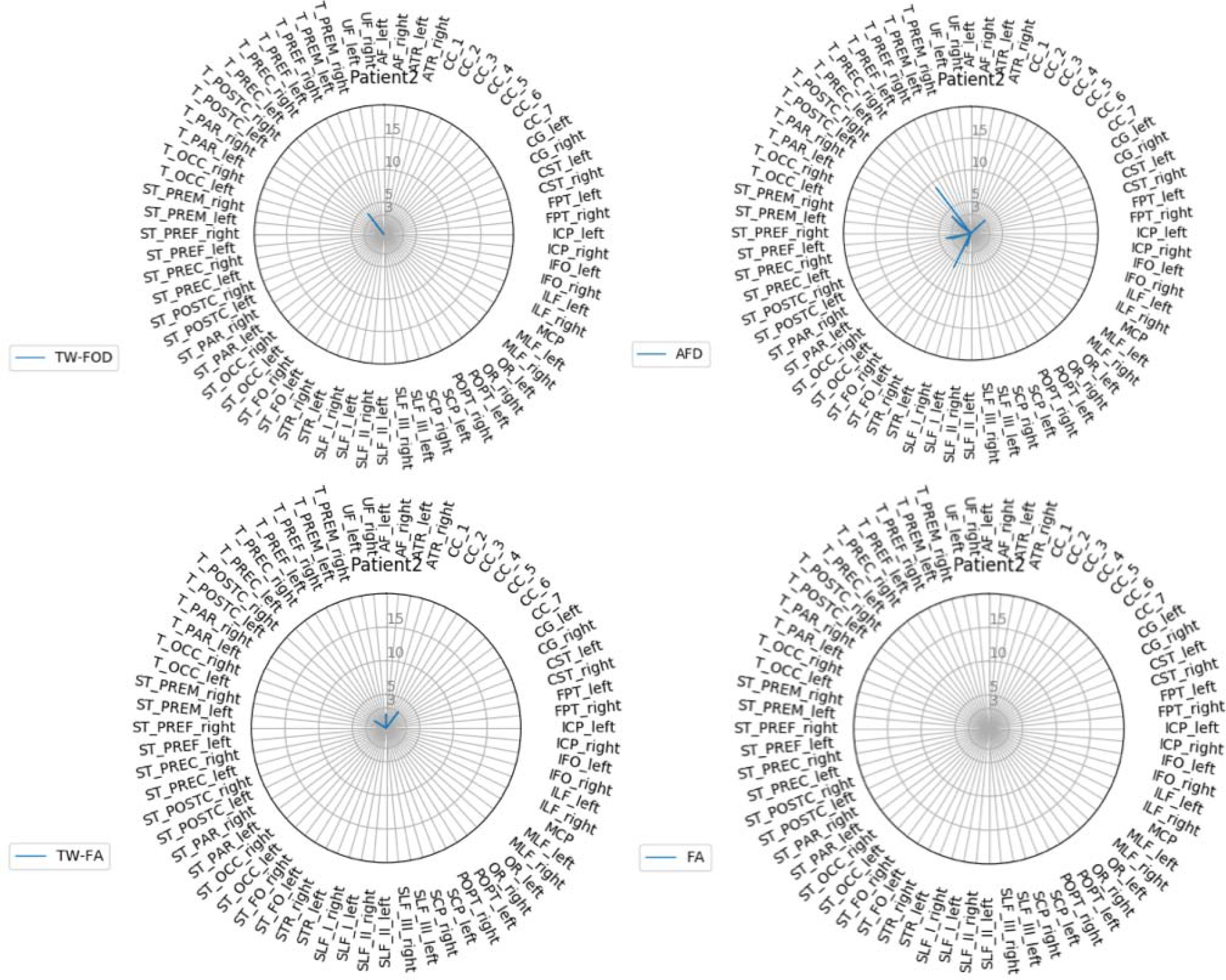
Radar plots of *TractLearn* results for patient 2, showing a low number of voxels presenting with significant altered Z scores in each bundle, as identified using *TractLearn* based on the session 1.

No morphological abnormalities were seen on morphological MRI sequences.

The back projections allow to identify the location of altered voxels within each bundle separately using the TW-FOD, TW-FA, bundle AFD and FA metrics, as illustrated in Figure 6 and 7 for two of the patients.

**Figure 6:**
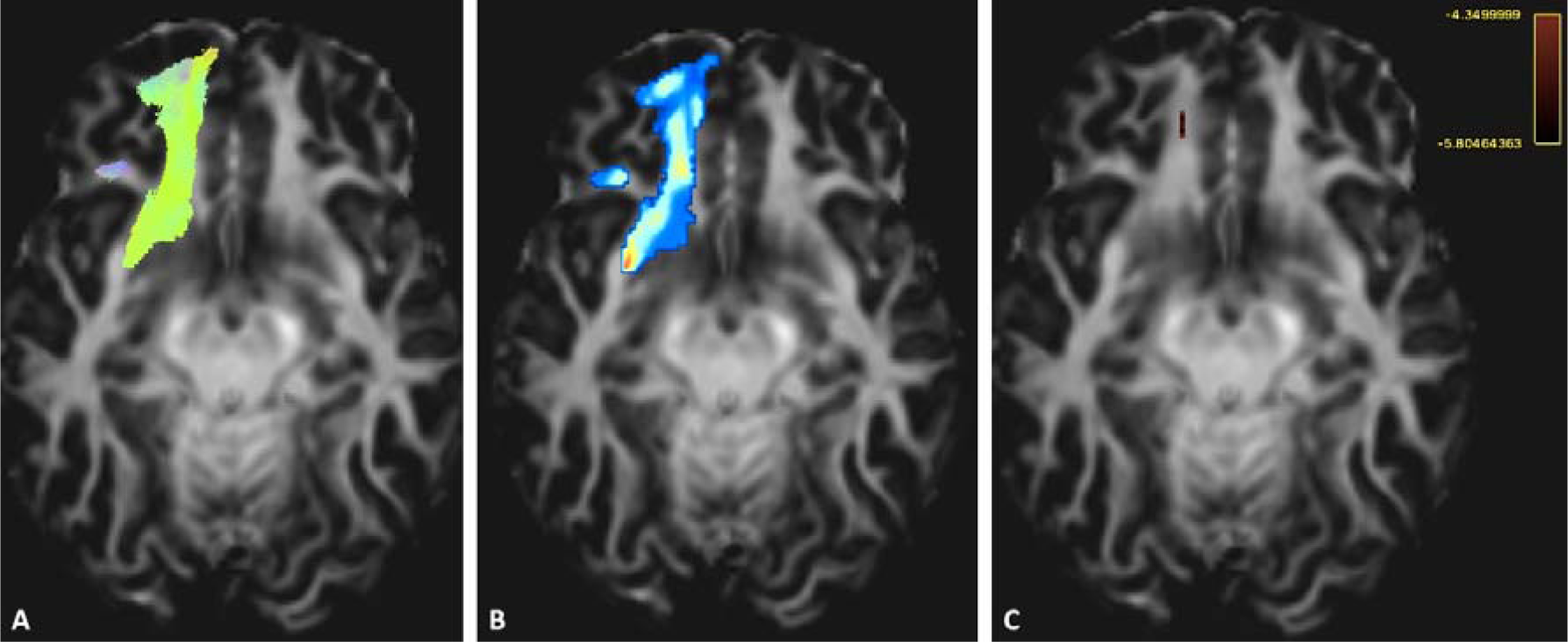
Back projection of the manifold residual in the template space, to localize the brain abnormality in Patient 1. Here is the example of the first part of the Striato-fronto-orbital (ST-FO right) with the track file identified by TractSeg (A), the track-weighted maps based on the TW-FOD contrast B) and the back projection of the Z Score (C). The Z-score threshold was set to -4,35; corresponding to a Bonferroni correction for a bundle of 7444 voxels. It has allowed the identification of voxels with significantly different TWI contrast, in comparison with the control group (here retest session).

**Figure 7:**
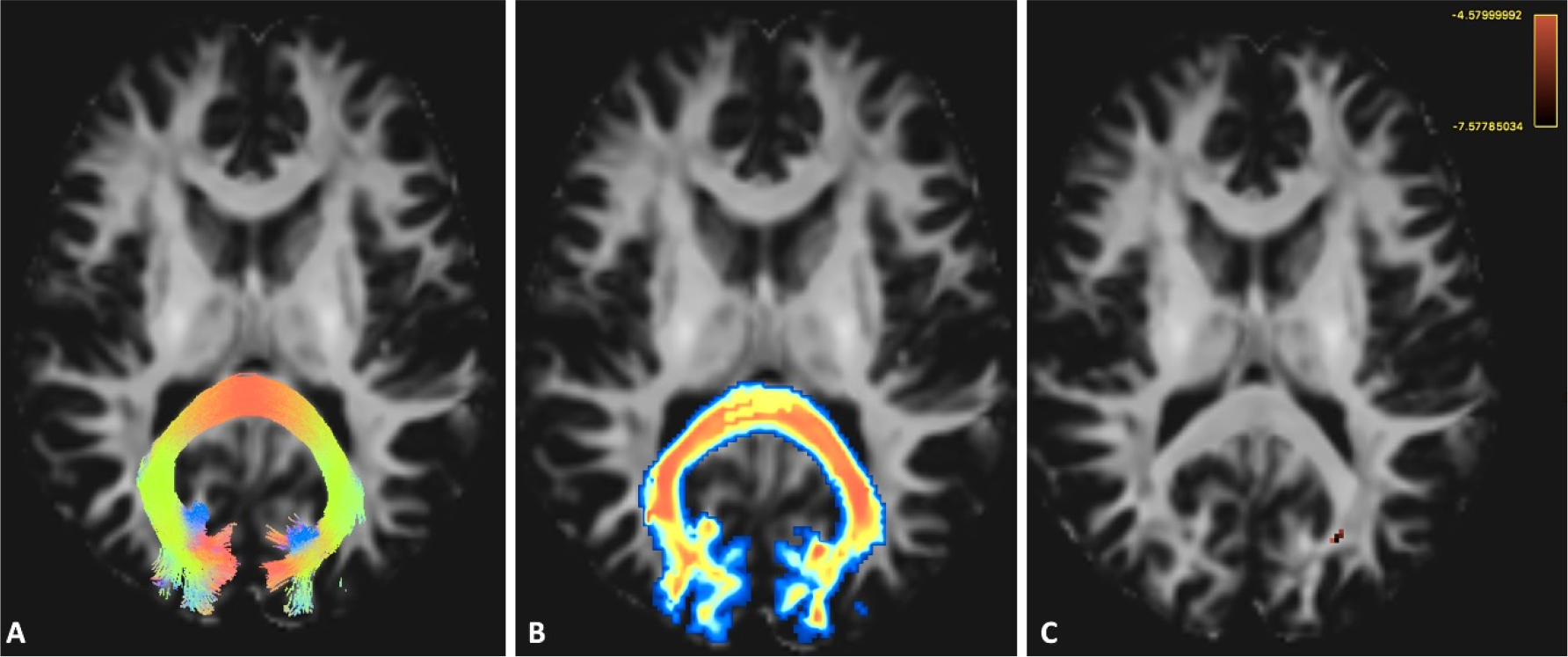
Another example from the first part of the corpus callosum part 7 (Patient 1) with the track file identified by TractSeg (A), the track-weighted maps based on the AFD contrast B) and the back projection of the Z Score (C). The Z score has allowed the identification of voxels with significantly different AFD contrast (p<0.05 Bonferroni corrected for colored voxels), in comparison with the control group. The Z-score threshold was set to - 4,73; corresponding to a Bonferroni correction for a bundle of 21303 voxels.

Using the Zero-one loss function for classification learning (i.e. to assign 0 to loss for a correct concordance between bundles with more than 5 altered voxels on the learned manifold built using the first MR session and bundles with more than 5 altered voxels on the learned manifold built using the second MR session: 0 is considered as a perfect score while 1 implies a total absence of concordance for all bundles), we have found scores as being 0.0029, 0.0088, 0.0147, 0.0029 for FA, TW-FA, AFD, TW-FOD, respectively (see Figure 8 for an example)

**Figure 8:**
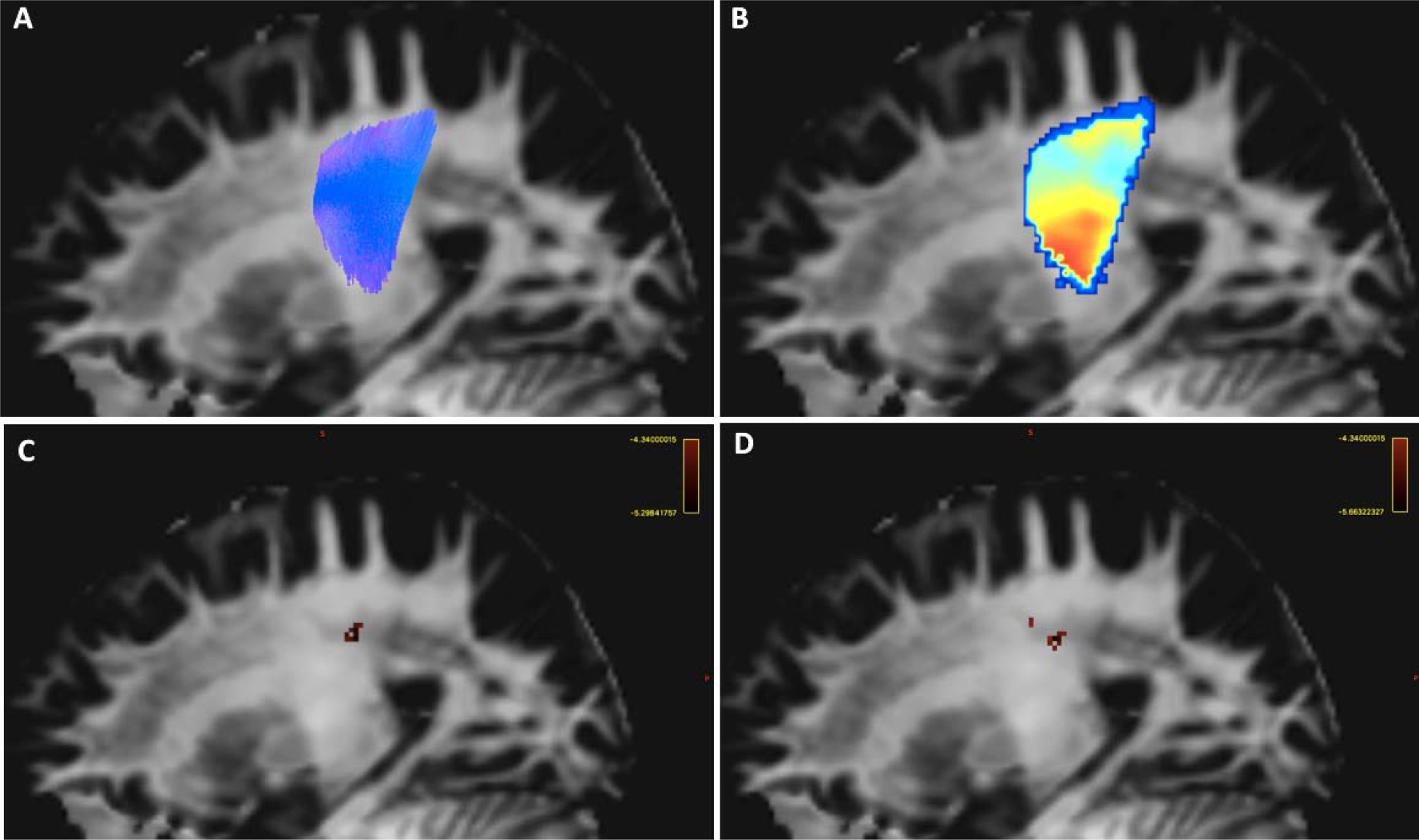
Example of the patient 4 left Superior thalamic radiation (A) and the subsequent track-weighted fractional anisotropy contrast (TW-FA, B). The image C illustrates the location of a local association of altered voxels (more than 15 voxels as identified using the radar plots) based on the test session manifold. The image D illustrates the altered voxels (here represented as blue voxels) based on Z-score calculated from the manifold of the retest session. The Z-score threshold was set to -4,34; corresponding to a Bonferroni correction for a bundle of 4100 voxels.

## DISCUSSION

We have proposed a unified framework for the quantitative analysis of properties of brain bundles by geodesic learning. *TractLearn*, a data-driven learning task, allows mapping between the high dimensional image domain and a reduced latent space of brain fascicles. In the patient group, *TractLearn* detected white matter abnormalities in a number of bundles; importantly, our results show that the identification of the abnormal bundles was reproducible, by comparing the number and location of abnormalities between a learned manifold built using the test session of controls and another learned manifold based on the retest session ~1 year later.

The injuries location identified by *TractLearn* (mainly located in the frontal lobes, occipital lobes and in the corpus callosum) are compatible with the known pathophysiology of mTBI (Delouche et al., 2016; Stokum et al., 2015) and were not visible on conventional MR sequences. In addition, we proposed a statistical framework having the capabilities to detect a potential interaction between voxel alteration through their representation in the reduced manifold space. As the voxels are not analyzed independently, we can theoretically identify subtle metrics variation in a given bundle. Finally, we proposed to use a method of back-projection from the highly sensitive low-dimensional manifold space to the MRI individual subject space, to visualize the physical location of the brain abnormalities.

For the examples shown in our study, we have used some selected illustrative DWI-related metrics (i.e. FA, bundle AFD, TW-FA and TW-FOD); it is however straightforward to extend the *TractLearn* framework to include other metrics, including exploiting the flexibility of the TWI approach, which allows to combine a tractogram with any other (even non-MRI) co-registered metric (Calamante, 2017). *TractLearn* only needs a collection of quantitative metrics for each bundle as input.

### 1. Benefits of manifold learning

A common problem in DWI studies is the difficulty to detect abnormalities at the single subject level (e.g. for precision medicine), and instead having to rely more commonly on group-based studies. The possibility to detect subtle fiber architecture alterations at the voxel level in individuals through direct analysis of multiple MRI contrasts requires advanced tools with the ability to manage high-dimensional vector space. Typical clinical studies often include a limited number of patients; the natural mathematical space of the quantitative values at the fascicle level then needs to be constrained to control for the mismatch between the number of samples (here the number of controls/patients) and the number of features (here the number of voxels in each bundle). This problem, which occurs in the context of HDLSS data, is known as the curse of dimensionality, and we therefore need to reduce the space, for example to be able to apply Z-score algorithms on quantitative analysis.

Dimensionality reduction seeks to produce a low dimensional representation of high dimensional data that preserves the original structure. Dimensionality reduction algorithms offer a solution to important problems in data science. They allow visualization of complex data and they can be applied as potential pre-processing step for machine learning. They are classically divided into two categories: those that seek to preserve the original distance structure within the data, and those that favor the preservation of local distances over global distance. The second category of algorithms includes for example t-SNE, the state-of-the-art in dimensionality reduction for visualization (Tenenbaum et al., 2000). The advantage of *UMAP* over t-SNE is a better preservation of the original structure (here the metrics coming from brain bundles) (McInnes et al., 2018) while being as powerful for visualization purpose.

We also proposed a generalization of the classical Z-score where *f(x)* corresponds to the mean value. Indeed, we have modeled the population by a regression function instead of a unique sample. In other words, all healthy controls do represent the population of reference instead of a unique individual. The first consequence is that each new studied subject in a previously learned manifold will be projected as close as possible to one of the control subjects (i.e. those with metrics values closest to the new subject) (see supplementary figure S1). In the classical Euclidean framework, there is no local mean as proposed but global mean, thus all tested subjects will have the same reference. This automatically leads to a higher standard deviation in metrics comparison, which in turn will hinder the possibility to detect quantitative abnormalities as the residual *ε* does contain the standard deviation added to the potential pathological effect.

Tilquin et al. (Tilquin et al., 2019) have already proposed to learn the manifold spanned by the normal controls using non-linear dimensionality reduction techniques. In their work, based on T1-weighted imaging, the image of a subject is projected on the control group manifold allowing a comparison of the reconstruction with the subject’s original neuroimaging data. The objective of these projection techniques is to detect abnormal patterns by way of statistical tests on the residuals. In this prior work, the importance of non-linear modeling of the manifold in the reduced-dimension subspace was highlighted, as well as robustness to abnormalities detection on a larger dataset, yet without retest session to assess the reliability, as we were able to include in our study.

Manifold learning has also been previously used in DWI. For example, the estimation of the embedded reduced space for DTI-based tensors has been previously described using Manifold Learning (Khurd et al., 2007). More recently, manifold learning has also been used on DWI to map the white matter fiber in a controlled subspace with an adapted model of fingerprint called “fiberprint” and on multimodal MRI acquisition including DWI data (Kumar et al., 2018, 2017); these studies have shown interesting relationship between compact fingerprint and genetic biomarkers on a large population data.

The possibility to back project the manifold information (here the Z-score testing) in the subject space help to precisely locate a disease process or pathophysiology in a patient-specific basis. Taken into account that the DWI data used here is consistent with a clinically suitable protocol (e.g. total acquisition time of DWI sequence <10 min), it can be anticipated that comparable level of sensitivity to detect brain alterations is possible with other similar acquisition protocols. It should be emphasized that patient-specific abnormalities were here detected either using the first learned manifold (built from the test session of the healthy controls) or using the second learned manifold (built from the retest session ~1 year later). The global effect on voxels metrics owing to the acquisition condition (test or retest) can be visualized in the reduced subspace. Regardless of the learned manifold chosen, the resulting number and location of abnormalities showed good correlation using the zero-one-loss test. It should be noted that Cohen’s Kappa should be avoided as performance measure due to the unbalanced nature of our dataset (Delgado and Tibau, 2019) (i.e. an important mismatch between the high number of bundles with no voxel abnormalities and the low number of altered bundles).

This suggests that, in our dataset, the local effect due to local axonal shear lesion or bleeding (associated with mTBI) is larger than the global effects related to the use of either the test or retest session to compute the learned manifold.

It could be speculated that this may also suggest that TractLearn might be more lenient to be able to detect local (disease related) effects over global (acquisition related) effects when using data from various acquisition or scanning protocols. This remains to be tested in future work, for example using datasets coming from multicenter studies.

### 2. Riemannian versus Euclidean statistical analysis

The GLM, based on a Euclidean framework, has been widely used in the medical and neurosciences communities, and has been implemented in popular software tools, such as Statistical Parametric Mapping (SPM, https://www.fil.ion.ucl.ac.uk/spm/). While being effective in the case of lowdimensional data comparison for group study, its main limitation relies on its inability to properly model the data distribution without using a smoothing step. The smoothing of Diffusion-weighted data usually decreases the standard deviation of all voxels metrics coming from the GLM, yet here we have shown that it leads to standard deviation much higher than in our framework (as illustrated in the supplemental material). It could be argued that smoothing still occurs within the TWI calculation; this however can be seen as a ‘smart-type’ (streamline-informed) smoothing (Calamante, 2017).

Imposing the Euclidean topology led to poor estimation because on the non-linear topology of the brain. Such inabilities to model quantitative data relying on complex topology have been demonstrated using dimensionality reduction by Tenenbaum (Tenenbaum et al., 2000), but also for comparing T1-weighted imaging anatomy (Miller, 2004) or high angular Diffusion-weighted data (Goh et al., 2011). In addition, using Riemannian distances allowed us to detect joint statistical variations in a group of voxels.

Here we have used *UMAP*, which relies on non-Gaussian and robust modeling, to capture the variability of the brain fascicles’ metrics. It should be noted that, even including a low sample data size for controls - with its inherent limitations to have potential anatomical variations displayed in the manifold, we were able to detect local bundle alteration in individual patients with mTBI.

### 3. The benefit of using TWI as a quantitative biomarker

TWI has proved to be powerful in mapping fascicles alterations in neurodegenerative diseases (Bozzali et al., 2011; Ziegler et al., 2014) or mTBI preclinical imaging (Tan et al., 2016), among others. The TWI contrast intrinsically contains the anatomical information of the fiber architecture (through the track-weighted averaging step), while also allowing a way to combine it with other local quantitative information (e.g. FA and FOD, in the current study). Numerous studies have proven the benefits of extracting FOD-related parameters for group studies, such as for example in the case of AFD in motor neuron disease (Raffelt et al., 2012b) or, more recently, Alzheimer’s disease (Mito et al., 2018). FA analyses, based on the tensor model, have been widely done, and they have allowed us to increase our understanding of numerous brain diseases, including traumatic disorders (Ilvesmäki et al., 2014) (Aoki et al., 2012), neurocognitive diseases (Bozzali et al., 2011) and in optic pathways studies (Bender et al., 2014; Mandelstam, 2012). These were some of the reasons for our choice of possible parameters to illustrate *TractLearn* in the current study.

Here, more brain bundles alterations were detected using the bundle AFD metric. Our work is, however, limited by the inability to validate our findings with pathological analyses, thus making difficult to compare the real added value of TractLearn over a more classical Euclidean approach. Future work is needed to address the sensitivity and specificity of the abnormalities detected by *TractLearn*.

As emphasized above, the track-weighted contrast uses a ‘smart’ smoothing effect (of the FOD amplitudes or FA values, in the current study) along the streamline. This work complements that of Willats et al. (Willats et al., 2014), which has demonstrated that the track-weighted contrast had good reproducibility in a sample of 8 subjects. Here we find high ICC values in 68 different brain regions of very different sizes in the context of a relatively large test-retest protocol (one year between sessions, instead of <2 weeks in the work of Willats et al). We also note that TW-FA was shown to be more reproducible than FA using a paired t-test on each bundle, consistent with the work of Willats et al. In contrast, we did not find a significant difference between TW-FOD and AFD ICC values. It should be noted however, that TW-FOD is not constructed based on the bundle AFD map and, therefore, their relationship is not as direct as the FA case (cf. TW-FA is constructed based on the FA map). Furthermore, bundle AFD mapping does also includes some smoothing along the bundle (related to the *afdconnectivity* command in MRtrix), and thus the benefits of increased reproducibility with the track-weighted approach in TWI is limited when compared with bundle AFD.

Interestingly, the Zero-one loss function results indicated that FA had higher concordance of detecting an abnormality than TW-FA. This might relate to the very focal abnormalities in the mTBI patients included in our small cohort, as then the streamline-specific smoothing involved with TW-FA could dilute the effect of the abnormality rather than amplify it (Calamante, 2017). Future work with a larger range of patient severities is required to fully characterize the benefits of TWI vs using DTI metrics directly.

### 4. Spatial coregistration

Most group DWI neuroimaging studies are registered to a common template. Here we have used a custom-made FOD template for robust coregistration of each individual FOD maps to the template, and thus obtain accurate matching between white matter structures (Raffelt et al., 2011); the resulting spatial transformation was applied to the TractSeg tractograms for each subject, to ensure correspondence between the identified bundles. This also ensures that the FODs and tracks lie on the same space, in order to correctly compute TW-FOD values (Calamante, 2017). However, we have here limited the anatomical variability of cortical termination by keeping only the top 80% voxels values of each bundle, based on the track-density contrast. Indeed, while the coregistration based on FOD symmetric diffeomorphic has allow to match major brain bundles, we have noticed that cortical variability made more difficult a perfect matching for the entire bundle. Absence of this step could potentially lead to false positive lesions on the bundles boundaries. In addition, as TractSeg tends to produce bundle overlaying (i.e some boundaries voxels can be linked to two different bundles), using a thresholding has allowed to precisely locate abnormalities.

### 5. Future perspectives

In medicine, researchers often raise a prior anatomical hypothesis on potential diseased brain bundles from which the quantitative analysis would allow to early detect the prodromal stages. Here we propose not to pass through this step to directly study a large part of the brain fascicle in a semi-supervised way (as the status of each point in the manifold is known before the calculation of the residual, i.e. whether the subject is a healthy control or a patient).

While *TractLearn* was here used for precision medicine (i.e. one vs. a healthy group), in general, it could be also applied on the more classical approach of group comparison studies, such as by replacing Z-score back projection with p-values back projection. The objective would be then to detect brain bundles alteration at the group level taking advantage on the Riemaniann framework. With the appropriate statistical models in hand, we may also regress the manifold data directly against one or more independent variables, for example clinical, biological or pathophysiological data.

Finally, while here we have used one scalar value per voxel for each manifold (e.g. a manifold for TW-FOD and a different one for TW-FA, etc.), it might be the case that some pathologies will not be able to be characterized just with one parameter. Similarly, it might be the case that higher order dimensions are needed to represent certain DWI features (e.g. using a manifold of the spherical harmonics amplitudes of the FODs rather than just the TW-FOD). The manifold framework should allow to decompose the DW signal by providing the same algorithm a collection of MRI metrics per voxel simultaneously.

### 6. Conclusion

We have presented *TractLearn*, a unified framework for brain fascicles quantitative analyses by geodesic learning. The possibility to detect abnormalities in individuals in the context of high-dimensional low sample size data hold promise for precision medicine.

## Data Availability

The code for TractLearn will be uploaded on Github on acceptance of a final version of the manuscript

## Acknowledgments

The computation server used benefitted from the ERATRANIRMA project, by Agence Nationale de la Recherche [grant ANR-12-EMMA-0056]. We would like to acknowledge the funding from the Alain Rahmouni grants of the French Society of Radiology. This work was also supported by funding from the National Health and Medical Research Council of Australia and the Australian Research Council.

ABBREVIATIONS
TWI: Track-weighted imaging
GLM: General linear model
HDLSS: High dimensional low sample size data
UMAP: Uniform Manifold Approximation and Projection

## Supplementary material

**Figure S1:**
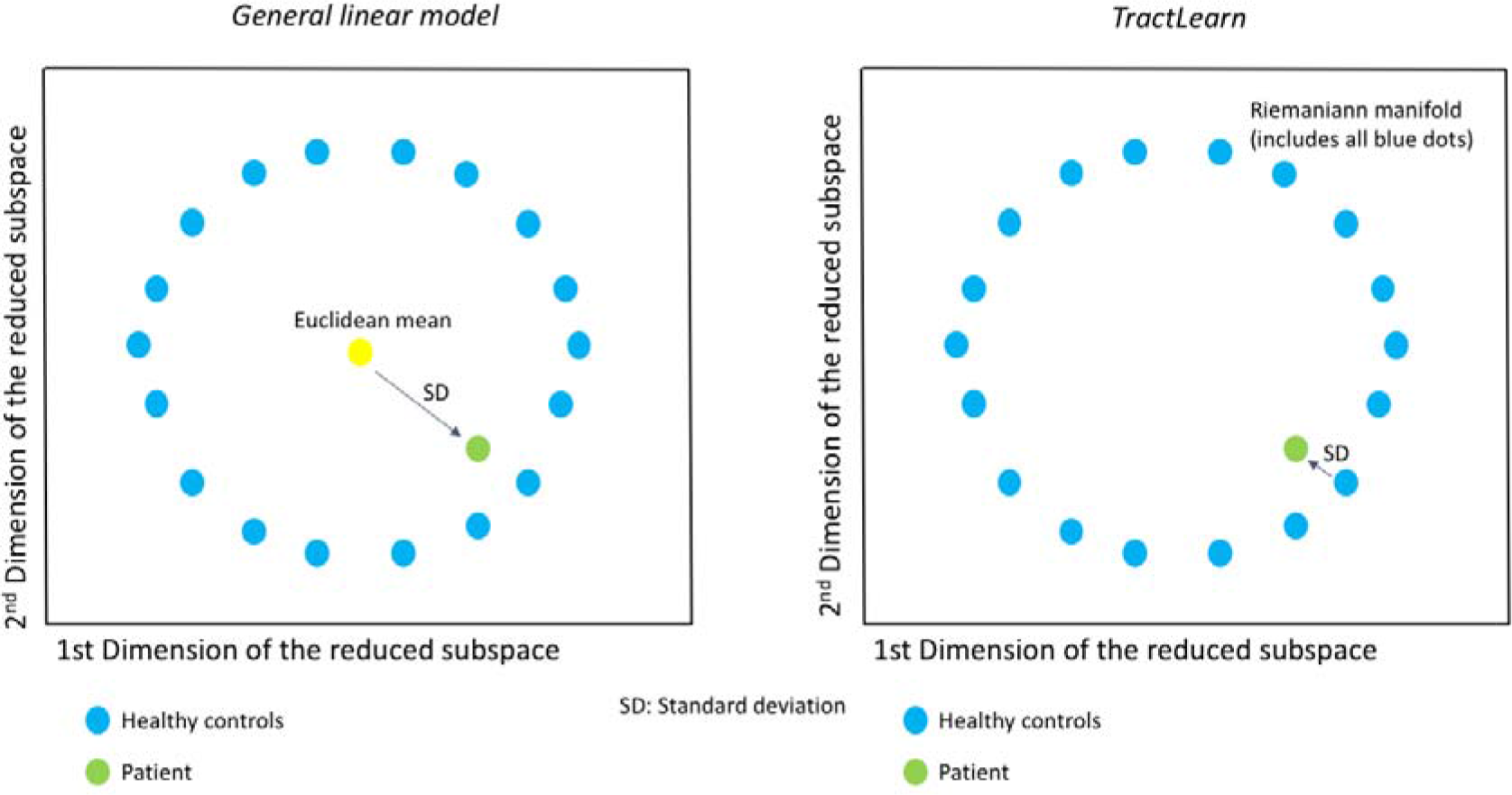
Toy example data in a reduced subspace, illustrating the comparison between the GLM and *TractLearn* healthy control population modeling. The SD is assumed to be smaller using *TractLearn* than using the GLM.

**Figure S2:**
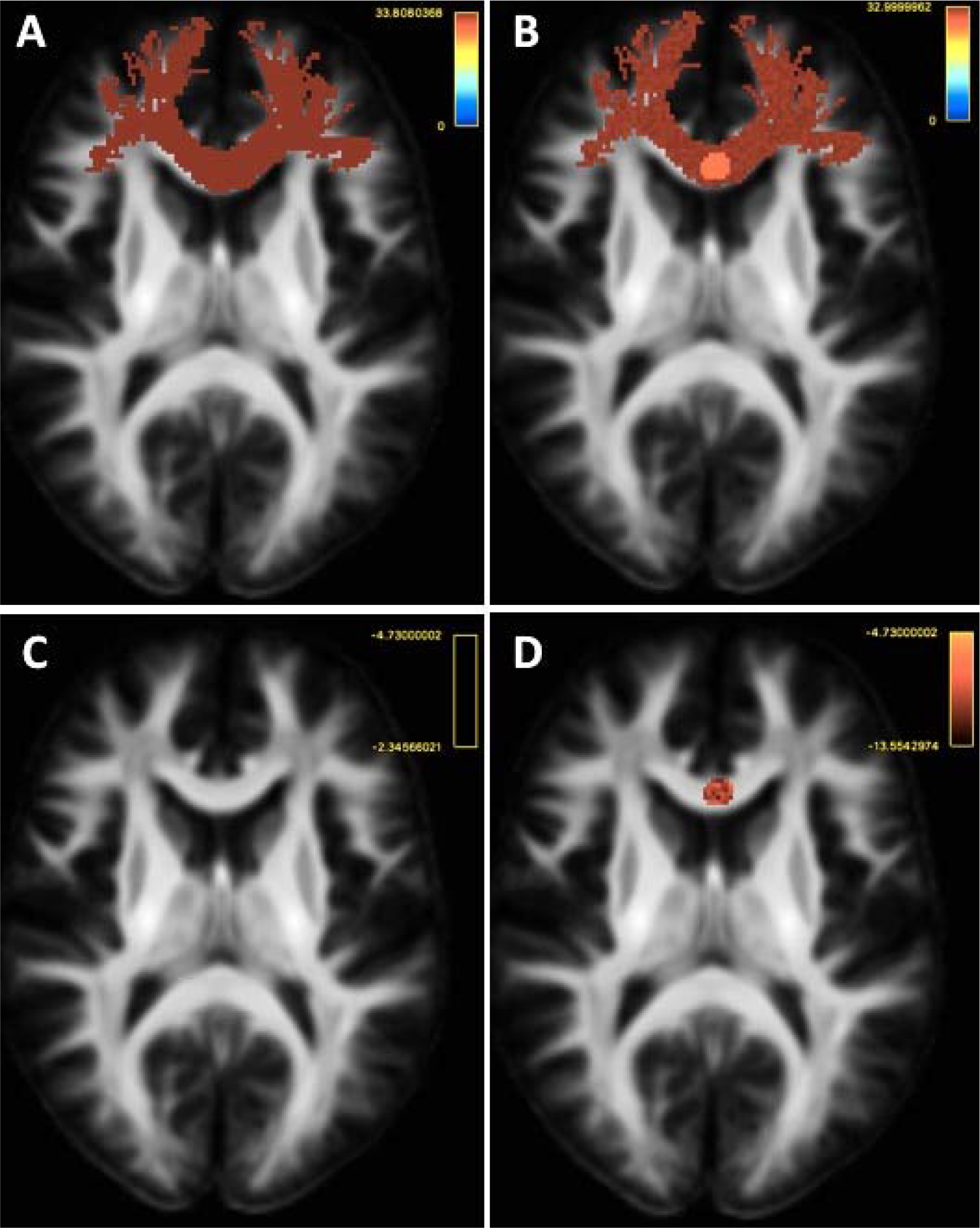
Illustration of the sensitivity of *TractLearn* to changes in the WM properties at the individual level using a toy simulated-example. A: Synthetic mean mask of CC2 coming from 50 healthy controls based on track-density imaging masks. Each synthetic track has a constant intensity following a Gaussian law N (33,3) to simulate anatomical inter-individual variability. In addition, a Gaussian noise N (0,1) was added for each control mask to simulate potential noise due to acquisition condition. This noise was reflected in the reduced subspaces by the absence of overlaying between the test and the retest session in each individual bundle. B: Example of a putative patient (PP) presenting a local lesion within CC2 (white arrow). In average, the putative patient keeps the same mean intensity than the control group. The lesion was simulated by a local decrease in voxels intensity (here the TDI value was decreased by -5). This value of -5 was chosen as closely similar to the inter-individual variability of the control, though higher than the acquisition noise, corresponding to a 17% decrease in intensity (as compared to the range of voxel intensity set between 0 and 33). C: Z scores obtained in the PP individual when compared to controls and based on the General Linear Model. The threshold was set to -4.73, corresponding to a Bonferroni correction adapted to a comparison of 47183 voxels, as contained in CC2 mask. No voxels are detected as deviant. D: TractLearn-based Z scores at the same threshold of -4.73. *TractLearn*, by finely taking account both inter- and intra-individual variability, was sensitive to a synthetic WM voxels alteration in a given bundle.

**Figure S3:**
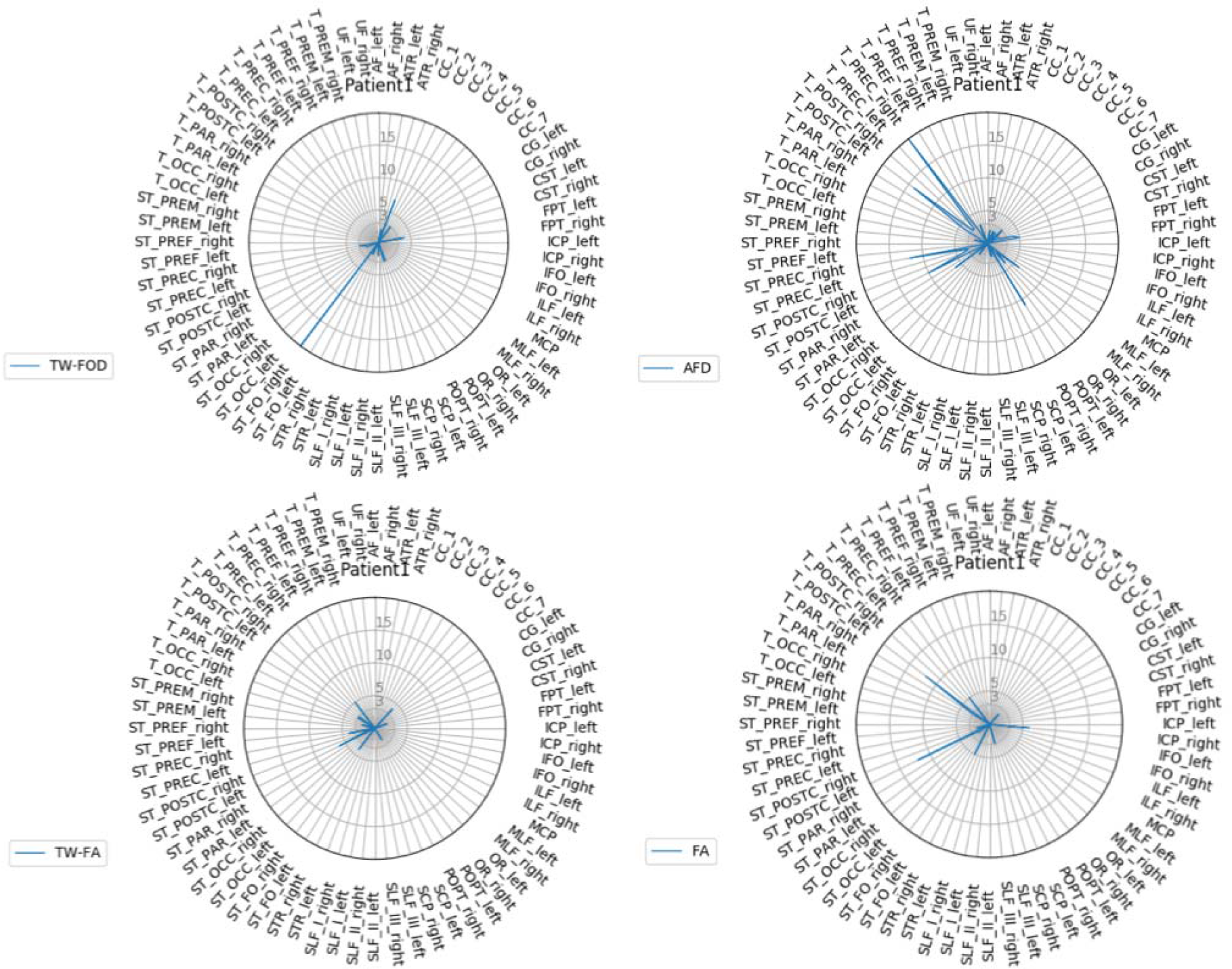
Radar plots of *TractLearn* results for patient 1, showing especially a high number of voxels presenting with significant altered Z scores in the right striato-fronto-orbital bundle using the TW-FOD metric and in the right thalamo-precentral bundle using the AFD metric.

**Figure S4:**
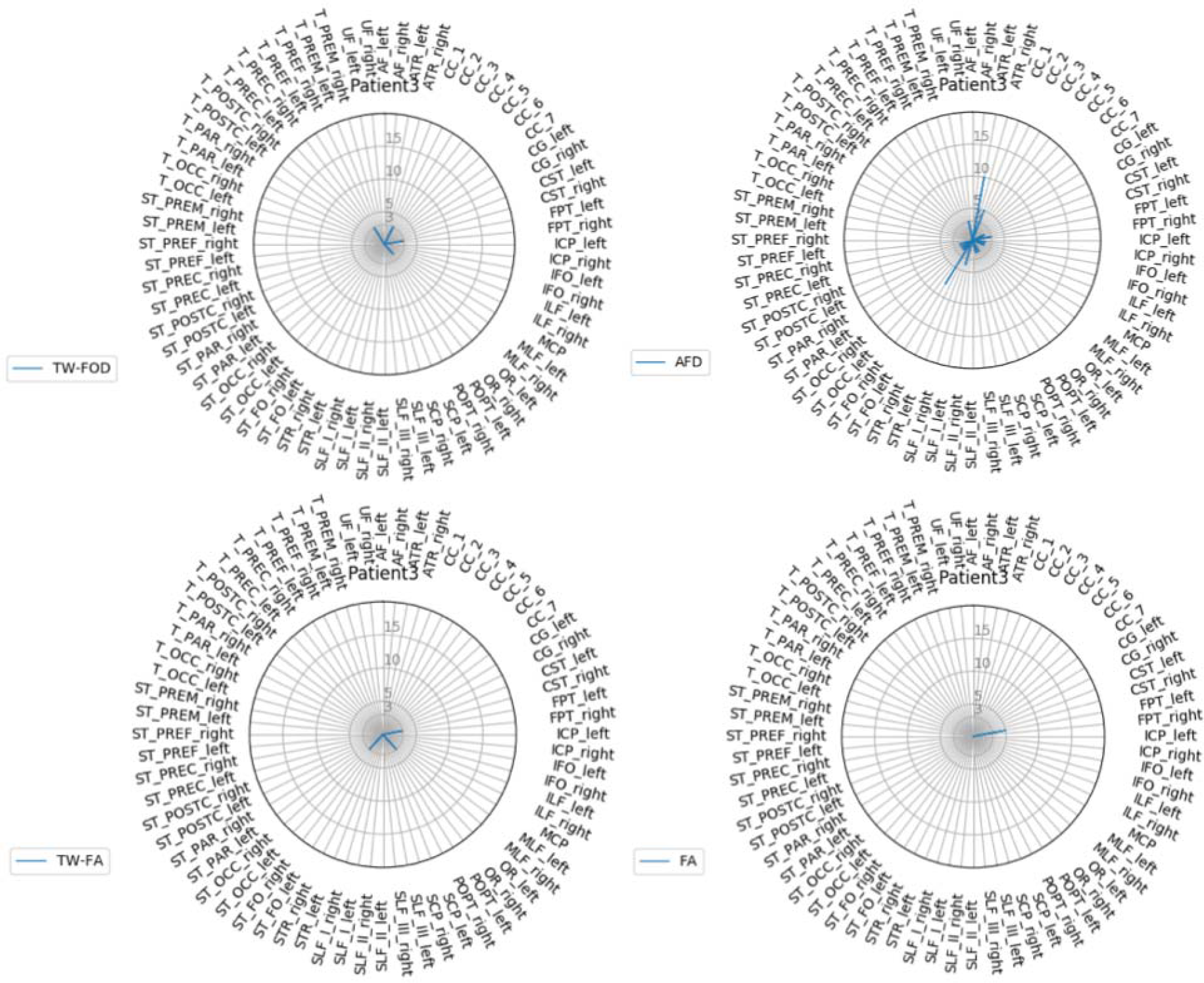
Radar plots of *TractLearn* results for patient 3, showing a low number of voxels presenting with significant altered Z scores, except in the anterior thalamic radiation using the AFD metric.

**Figure S5:**
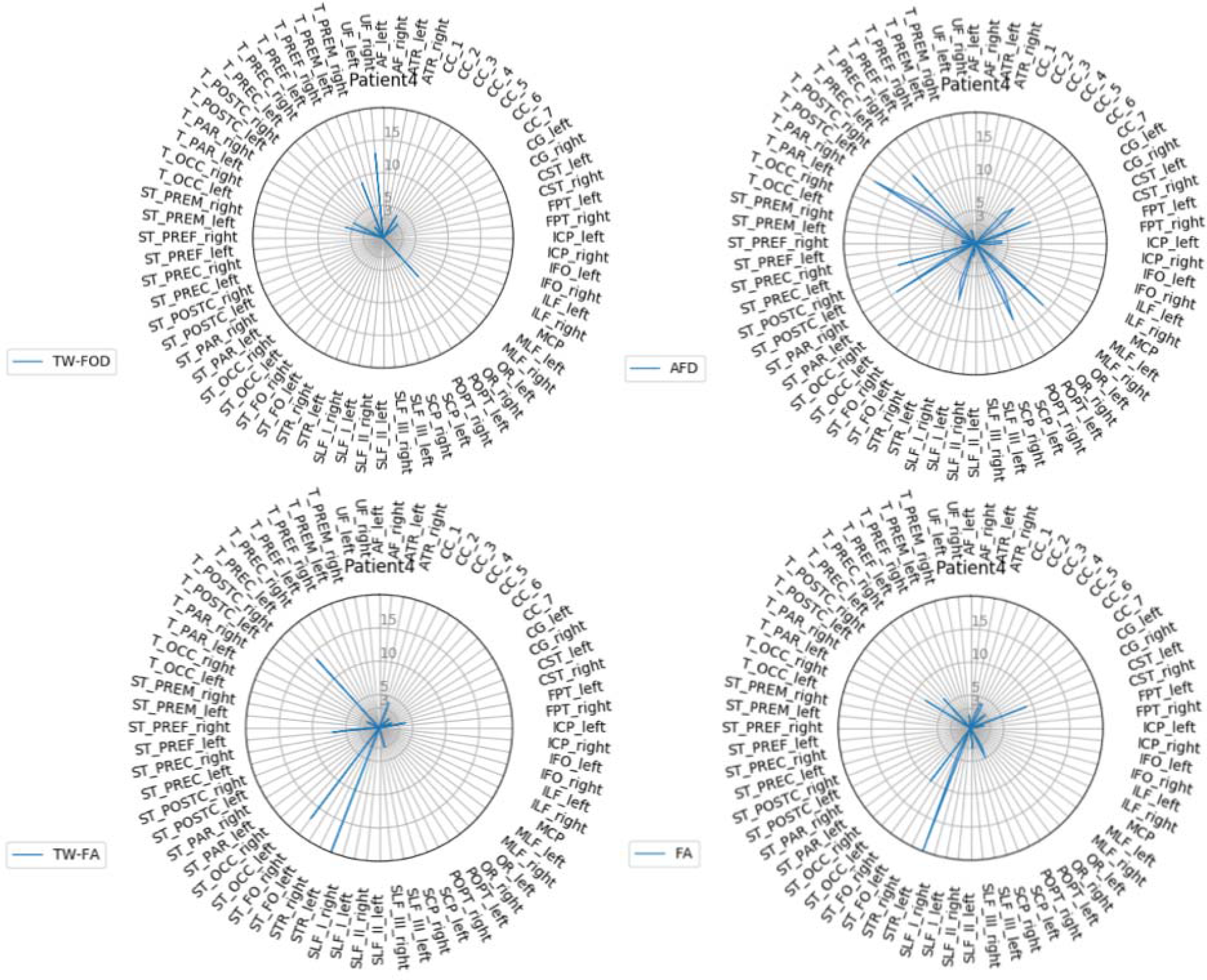
Radar plots of *TractLearn* results for patient 4, showing especially a high number of voxels presenting with significant altered Z scores in the left superior thalamic radiation using both the TW-FA and FA metrics.

## Footnotes

1 In MRtrix, *afdconnectivity*can be used to compute a map of the AFD estimated for each voxel, based on the sum of the FOD lobes aligned with the direction of the streamlines, divided by the streamline length (see https://mrtrix.readthedocs.io/en/latest/concepts/afd_connectivity.html for more details).

